# Eating Disorders and Parkinson’s Disease - 1: Comorbidities, Neurobiology and Family History

**DOI:** 10.64898/2026.01.03.25342594

**Authors:** Angeline Krueger, Andrew W Bergen, Irene Litvan, Vincent Filoteo, Carolina Makowski, Susan Murray, Karlee McGlone, Michael Garvin, Andi Drouin, Alyssa Kauffman, Caroline Steele, Walter H Kaye

## Abstract

**Objective:** In response to a clinical observation of an Anorexia Nervosa (AN) patient with family history of Parkinson’s Disease (FHoPD), and evidence of similarities in dopamine function, personality, anxiety and weight loss symptoms between AN and PD, we completed a pilot study to estimate FHoPD in families of those with eating disorders (EDs).

**Method:** We ascertained FHoPD among ED patients and community participants, and estimated relative risks (RRs) for AN, Bulimia Nervosa (BN) and Binge Eating Disorder (BED).

**Results:** We observed increased FHoPD among patients and community participants (N=482) meeting criteria for ED diagnoses compared to community participants (N=394) without an ED diagnosis. For AN, FHoPD prevalence was 6.6% vs 3.4%, χ^2^=4.638, p=.031, RR=1.935, 95%CI=1.012-3.768. For BN, FHoPD prevalence was 7.4% vs 3.4%, χ^2^=4.941, p=.026, RR=2.169, 95%CI=1.023-4.620. For BED, FHoPD prevalence was 13.3% vs 3.4%, χ^2^=6.953, p=.008, RR=3.886, 95%CI=1.108-11.524.

**Conclusions:** EDs are associated with an elevated FHoPD. Translational analyses leveraging disorder-specific research resources may benefit our understanding of the genetics and neuroscience of both disorders.

**Highlights:**

- AN and PD share premorbid anxiety, harm avoidance and dopaminergic dysfunction.
- FHoPD RRs are two-fold for AN and Bulimia Nervosa (BN) and four-fold for Binge Eating Disorder (BED) in a sample of treatment seeking and community participants.
- ED and PD familiality, and advances in PD and ED genetics and neuroscience research provide opportunities for cross-disorder research.

## Introduction and Aims

Anorexia nervosa (AN) is a complex neuropsychiatric-metabolic eating disorder (ED) with a strong genetic component (Watson et al. 2019; Bulik et al. 2022), and substantial morbidity and mortality in part due to limited effectiveness of treatments (Kaye and Bulik 2021). A better understanding of the genetic and biological mechanisms contributing to the neurobiology of AN may assist in the identification of novel and effective treatment targets.

When a patient with AN mentioned her father had developed Parkinson’s Disease (PD), and after confirmation that PD occurred in families of other individuals with AN, several clinical, neurobiological and genetic (Smeland et al. 2025) characteristics supported a possible cross-disorder relationship. PD is a movement disorder with complex genetic and environmental interplay, with rare juvenile, uncommon early onset, and common late onset forms with Mendelian genes, polygenes, and environmental and age-related risk factors (Trevisan et al. 2024; Ben-Shlomo et al. 2024). While PD may appear unrelated to the EDs given substantial differences in presentation and course, we review common characteristics that support the possibility of shared genetic vulnerability.

It is well known that AN is associated with extreme weight loss (Hebebrand et al. 2025). Half of PD patients experience unintentional weight loss during their illness, associated with nigrostriatal dopamine degeneration (Pak et al. 2018).

PD is associated with decreased CSF homovanillic acid (HVA) levels, the major metabolite of brain dopamine (DA), reflecting DA depletion in the nigrostriatal system (Jiménez-Jiménez et al. 2014). Individuals with AN who have been weight restored have reduced CSF HVA comparable in magnitude to PD (Kaye et al. 1999). DA neurons innervate striatal pathways, and altered striatal function has been found in individuals with AN (Fladung et al. 2013; Foerde et al. 2015; Kaye et al. 2020).

Individuals with AN and related EDs (e.g. bulimia nervosa, BN) tend to be highly anxious; and have a high prevalence of anxiety disorders such as Obsessive Compulsive Disorder (OCD) (Meier et al. 2015; Kaye et al. 2004). Individuals with AN often develop anxiety disorders in childhood before AN onset (Marzola et al. 2019; Deep et al. 1995). Anxiety is common in PD (Overton and Coizet 2020), occurring in at least 30% of patients (Jacob et al. 2010) and frequently presents before motor symptoms occur (Weisskopf et al. 2003). Anxiety in PD does not represent a response to the difficulties of adapting to the diagnosis or motor consequences of PD but is related to the neurobiological underpinnings of PD (Overton and Coizet 2020). In sum, anxiety occurs before the onset of PD and AN, suggesting that anxiety may be a risk factor for both.

Other commonalities between AN and PD include enhanced sensitivity to punishment (Jonker et al. 2022; Frank et al. 2004), as well as elevated harm avoidance (HA), found in individuals who are ill and recovered from AN (Atiye et al. 2015; Harrison et al. 2011) as well as PD (Santangelo et al. 2018). HA is a biologically informed conceptualization of trait anxiety (Cloninger et al. 1993) that reflects the heritable tendency towards behavioral inhibition to avoid punishment, loss, or frustrating non-reward. HA in AN and PD is associated with dopamine D2/D3 receptor binding in similar brain regions – the dorsal caudate (Kaasinen et al. 2001; Frank et al. 2005; Bailer et al. 2013).

We carried out a preliminary study to investigate the possibility of increased FHoPD in a relatively large sample of individuals with and without an ED diagnosis, and to provide evidence and rationale for whether a larger funded study is warranted. Because there is evidence for dopamine disturbances in binge eating (Yu et al. 2022), we included a broad range of ED subjects in this survey.

## Methods

All study procedures were approved by the University of California San Diego Institutional Review Board.

We included two groups of participants. Group A consisted of female and male individuals who met criteria for a DSM-5 ED and who were admitted for treatment to the UCSD Eating Disorders Center for Treatment and Research (EDCTR) Before receiving treatment in the EDCTR, prospective patients completed an intake assessment with a trained masters-level intake clinician. This included asking individuals with EDs who inquired about treatment and research about their family history of psychiatric and medical illness. In addition, their families were asked about any family history of PD (FHoPD). All research screenings were conducted by undergraduate-level trained research assistants. Group A consisted of 483 patients: 359 with AN (Restricting type, Binge-eating/purging type, or atypical AN), 98 with BN and 26 with Binge Eating Disorder (BED).

Group B consisted of two groups of community members: healthy control women, and women who met criteria for an ED diagnosis. We assessed prospective community research participants for FHoPD who were undergoing general study screeners to determine eligibility for various research studies conducted through the Eating Disorders Research Group. All community research assessments were conducted by undergraduate-level trained research assistants. As part of the screening process, relevant information was collected pertaining to eating disorder diagnosis, weight, and psychiatric and medical history. Group B consisted of 724 community members: 408 healthy women without a history of an ED and 244 with an ED (123 with an AN-Spectrum disorder, 117 with BN, and 4 with BED).

## Results

Of 727 individuals with an ED (483 Group A patients and 244 Group B community members), 52 (7.2%) individuals said that they had a relative with PD (**Table 1**). This was significantly more than the 14 (3.4%) of 408 healthy control women without an ED who said they had a relative with PD (p=0.01) (**Table 1**). Individuals with AN, BN or BED diagnoses endorsed a FHoPD at rates (6.6%, 7.4% and 13.3%) that exceeded the healthy women (3.4%) FHoPD rate (**Table 1**). Estimated RRs of a FHoPD in individuals fulfilling criteria for AN, BN and BED were two-fold (*p*=.031), two-fold (*p*=.026) and four-fold (p=.008), respectively (**Table 1**), but did not differ significantly among ED subgroups (*p*-values>.164) (**Table 2**). FHoPD rates were similar across individuals with an ED whether treatment-seeking or research-seeking (*p*=0.550) (**Table 3**).

**Table 1.**
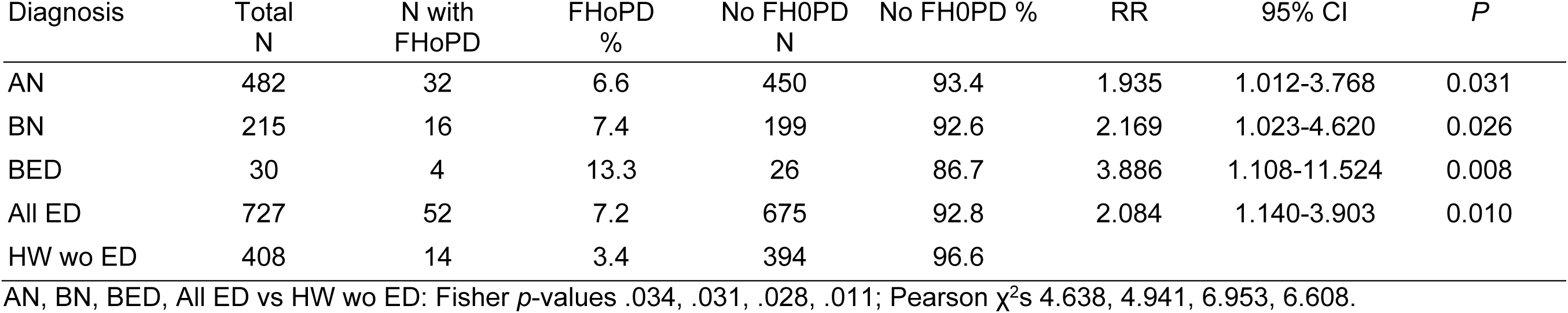
Relative Risk (RR) of FHoPD by ED diagnostic group compared to healthy women (HW) after combining Groups A and B.

**Table 2.** Comparing rates of FHoPD in 3 ED diagnostic groups, pairwise.

| ED subgroup comparison | ED1 diagnosis | ED1 N | ED1 FHoPD | ED1 FHoPD % | ED2 diagnosis | ED2 N | ED2 FHoPD | ED2 FHoPD % | *P* |
| --- | --- | --- | --- | --- | --- | --- | --- | --- | --- |
| AN vs BN | AN | 482 | 32 | 6.6% | BN | 215 | 16 | 7.4% | 0.699 |
| AN vs BED | AN | 482 | 32 | 6.6% | BED | 30 | 4 | 13.3% | 0.164 |
| BN vs BED | BN | 215 | 16 | 7.4% | BED | 30 | 4 | 13.3% | 0.270 |

AN vs BN, AN vs BED, BN vs BED: Fisher *p*-values 0.746, 0.151, 0.282; Pearson $\chi^2$ s 0.149, 1.936, and 1.219.

**Table 3.** Comparing rates of FHoPD in patients and community members with ED.

| ED subgroup | N* | FHoPD* | FHoPD%* | No FHoPD* | No FHoPD%* | *P* |
| --- | --- | --- | --- | --- | --- | --- |
| EDCTR | 483 | 30 | 6.2% | 453 | 93.8% | 0.550 |
| Community | 244 | 18 | 7.5% | 226 | 92.5% |  |

EDCTR vs Community: Fisher *p*-value=0.532, Pearson $\chi^2$ =0.357.

## Discussion

The current study provides preliminary evidence for significant genetic links between ED and PD through examination of family history. Observed FHoPD RRs (two- to four-fold) were similar to reported RRs of AN within families of individuals with AN (Steinhausen et al. 2015) and of PD within families of individuals with PD (Thacker and Ascherio 2008), but did not differ between ED diagnostic groups and did not differ between patients and community participants with ED. As discussed below, this paper proposes that leveraging advances in PD genetics and neurobiology (given PD’s greater research resources) may improve our understanding of AN neurobiology, as well as suggest translational research for both disorders.

### Anxiety, AN and PD

Meier proposed that anxiety may share etiological mechanisms with AN, and/or anxiety represents one developmental pathway to developing AN (Meier et al. 2015). Moreover, the presence of anxiety has been shown to increase the severity of AN (Lilenfeld et al. 1998; Dellava et al. 2010; Raney et al. 2008; Bloss et al. 2011). Population-based studies find increased rate ratios of PD in those with existing and late-onset anxiety diagnoses (Svensson et al. 2016; Bazo-Alvarez et al. 2024), especially in patients with OCD (Liou et al. 2022). While individuals with ED and with PD suffer from anxiety disorders,there has been limited progress in developing more effective treatments for anxiety.

### DA receptors, anxiety and AN

Recent research supports dopamine systems as a critical regulator of aversive learning and memory (Castell et al. 2024); DA neurons affect the salience of stimuli predicting rewarding or aversive outcomes and this is critical for adaptive behaviors, possibly including the HA construct. Perez found that amygdaloid DA D2 receptors (DAD2R) play an important role in the modulation of unconditioned fear and anxiety (Perez de la Mora et al. 2012), while Tsutui-Kimura found that DAD1R and DAD2R interactions in the tail of the striatum regulates threat avoidance (Tsutsui-Kimura et al. 2025). Castell found that mice in which DA D2 was selectively deleted had an enhanced excitatory and inhibitory response in ventral tegmental area DA neurons to aversive stimuli, suggesting that DAD2Rs act as a critical regulatory brake. Finally, in recovered AN, increased self-reported anxiety was positively associated with increased endogenous dorsal caudate DA concentrations (Bailer et al. 2017). In comparison in healthy control women,endogenous DA in the ventral striatum tends to be associated with euphoria (Bailer et al. 2017). This finding could explain why food-related DA release is associated with anxiety in AN, whereas feeding is pleasurable in healthy participants.

### DA receptors and anxiety

Replicated studies show that unmedicated individuals with an ED who have recovered have dorsal striatal DA D2/D3 receptor binding (DA D2/D3R) positively correlated with harm avoidance (Frank et al. 2005; Bailer et al. 2013). In PD, endogenous DA function is positively associated with elevated HA in the right caudate (Kaasinen et al. 2001), a homologous area to the dorsal striatum. Identification of similar neurobiological mechanisms (e.g., HA associated with DAD2R activity) in AN and PD could greatly advance our mechanistic understanding and stimulate developing more effective treatments for both disorders. Moreover, comparative studies in PD may raise consideration of new insights into the genetics, neural pathways, and molecular biology of ED. For example, Overton (Overton and Coizet 2020) proposed that anxiety in PD is related to an elevated response to threat-related stimuli due to a hyper-responsive superior colliculus (SC), the “front-end” of a sensory system in the brain (SC-pulvinar-amygdala) that modulates rapid reflexive threat detection and becomes hyper-responsive to sensory stimuli following dopamine denervation of the striatum in PD.

We hypothesize that post-synaptic DAD2R becomes upregulated to compensate for diminished endogenous dopamine in both AN and PD. Montardy (Montardy et al. 2022) stated that the superior colliculus (SC) is known for a role in defensive behaviors to visual threats, and that optogenetic activation of SC DA D2 neurons trigger defensive behaviors whereas inhibition of these neurons decreases defensive behaviors. The SC receives integrated information from higher-order associations and prefrontal cortical areas (anterior cingulate, retrosplenial, and posterior parietal cortices), as well as prefrontal regions (infralimbic, prelimbic, and orbitofrontal cortices). These regions integrate spatial, auditory, visual, and somatosensory information to the SC. Melleu (Melleu and Canteras 2024) described pathways by which the SC and basal ganglia interact and constitute part of neural networks that process signals regarding navigation, memory, and spatial orientations as well as interoceptive and emotional components from limbic structures. In summary, the wealth of data exploring the pathophysiology and genetics of PD may prove to be valuable in generating new insights into ED and may accelerate new insights into the pathogenesis and symptom complexes of both disorders.

### DA function, ED, PD, and genetics

While disturbances of DA function occur in both disorders, there is little evidence that genetic alterations occur within DA-related genes (receptors, transporter, or metabolic enzymes). Rather, DA disturbances in PD appear to be a consequence of disturbances of genes or proteins that support the necessary cellular metabolic pathways, including energy metabolism, protein degradation, organelle autophagy and synaptic transmission (Ramesh and Arachchige 2023). Three decades of PD gene discovery encompassing the linkage and genome-wide eras have resulted in hundreds of genes associated with PD. Defining molecular mechanisms in ED may be an area where insights from PD genetics may be instructive. Relatively little is known about the pathophysiology of ED in comparison to what is known about PD. If disturbances of DA neurotransmitter function are a consequence of disturbances of genes involved in neural structure and function, studies comparing the molecular biology of these neural pathways in ED and PD may help identify mechanisms and result in more effective treatments for dysfunction in both disorders.

Bergen and Makowski (Bergen, Andrew W et al., n.d.) found AN and PD jointly associated at rs1352420, a SNP in a region with genome-wide significant findings in AN and PD (Watson et al. 2019; Chang et al. 2017). This SNP is functionally linked to 40 genes in the complex chr3p21.31 region, is located in *IP6K2* and is adjacent to *NCKIPSD*. The inositol pathway was linked to PD using genome-wide PD findings (Yu et al. 2024) and NCKIPSD maintains dendritic spines and modulates synaptic activity in neurons (Cho et al. 2013). It may be that disturbances of neurotransmitter function in AN and PD are a consequence of disturbances of genes involved in neural structure and function (Murray et al. 2023). Thus studies comparing the molecular biology of these neural pathways in ED and PD may help identify biological mechanisms and result in more effective treatments for neurotransmitter dysfunction in both disorders.

### Limitations

The questionnaire administered to participants did not enumerate or directly interview relatives. The clinical study was performed at a single site in the United States, limiting generalizability. This study was a pilot proof of concept study; the lack of funding limited the resources available to gather more data on participants or investigate a larger sample of subtypes such as BED.

## Conclusions

This pilot study demonstrates significant familiality between ED (AN, BN and BED) and PD. This is the first description of increased FHoPD in families of AN probands, warranting further investigation. Since only a minority of individuals with ED have a FHoPD, these findings may be pertinent for only a fraction of individuals with ED. Still, it is possible that these data will provide new insights into the neurobiological mechanisms of ED.

We are not aware, and have never observed the development of PD in a patient with an ED. However, ED can progress into a chronic and disabling disorder for some individuals. It is not known whether a family history of PD may result in a more severe course of ED. However, we observed several individuals who have had excellent long-term recovery had relatives with PD, providing preliminary reassurance that a FHoPD does not increase vulnerability for a more severe ED or the onset of PD.

## Declarations

## Acknowledgments

We thank patients, community members and Eating Disorders Center for Treatment and Research staff for their participation and contributions to this study.

## Data availability statement

Available clinical data are summarized in Tables; contact the corresponding author with questions about data.

## Ethics approval statement

Ethical approval was granted by the UCSD Institutional Review Board.

## Permission to reproduce material from other sources

All sources of estimates and findings were cited.

